# Fever, Diarrhea, and Severe Disease Correlate with High Persistent Antibody Levels against SARS-CoV-2

**DOI:** 10.1101/2021.01.05.21249240

**Authors:** Maya F. Amjadi, Sarah E. O’Connell, Tammy Armbrust, Aisha M. Mergaert, Sandeep R. Narpala, Peter J. Halfmann, S. Janna Bashar, Christopher R. Glover, Anna S. Heffron, Alison Taylor, Britta Flach, David H. O’Connor, Yoshihiro Kawaoka, Adrian B. McDermott, Ajay K. Sethi, Miriam A. Shelef

## Abstract

Lasting immunity will be critical for overcoming the coronavirus disease 2019 (COVID-19) pandemic caused by severe acute respiratory syndrome coronavirus 2 (SARS-CoV-2). However, factors that drive the development of high titers of anti-SARS-CoV-2 antibodies and how long those antibodies persist remain unclear. Our objective was to comprehensively evaluate anti-SARS-CoV-2 antibodies in a clinically diverse COVID-19 convalescent cohort at defined time points to determine if anti-SARS-CoV-2 antibodies persist and to identify clinical and demographic factors that correlate with high titers. Using a novel multiplex assay to quantify IgG against four SARS-CoV-2 antigens, a receptor binding domain-angiotensin converting enzyme 2 inhibition assay, and a SARS-CoV-2 neutralization assay, we found that 98% of COVID-19 convalescent subjects had anti-SARS-CoV-2 antibodies five weeks after symptom resolution (n=113). Further, antibody levels did not decline three months after symptom resolution (n=79). As expected, greater disease severity, older age, male sex, obesity, and higher Charlson Comorbidity Index score correlated with increased anti-SARS-CoV-2 antibody levels. We demonstrated for the first time that COVID-19 symptoms, namely fever, abdominal pain, diarrhea and low appetite, correlated consistently with higher anti-SARS-CoV-2 antibody levels. Our results provide new insights into the development and persistence of anti-SARS-CoV-2 antibodies.

## INTRODUCTION

Severe acute respiratory syndrome coronavirus 2 (SARS-CoV-2) was isolated in January 2020 after an outbreak of 44 cases of pneumonia in Wuhan City, China (1). SARS-CoV-2 causes coronavirus disease 2019 (COVID-19), which ranges from no symptoms to a flu-like illness to severe respiratory distress syndrome and death (2). As of December 2020, there have been over 78 million cases worldwide and over 1.7 million deaths (3), with devastating effects on health, economies, and societies (4).

Lasting immunity, often estimated by persistent antibodies, will be critical for overcoming the COVID-19 pandemic, but our understanding of the development of persistent anti-SARS-CoV-2 antibodies is still emerging. In severe acute respiratory syndrome (SARS), caused by the related SARS-associated coronavirus (SARS-CoV), anti-viral antibodies persist in the majority of patients for at least 3 years after disease (5-7). Less is known about SARS-CoV-2, but a few reports suggest that immunity may last at least 6 months (8-10). However, other reports (11-14) suggest that antibodies against SARS-CoV-2 can decline in the first few months (15), with some patients becoming seronegative (16, 17). This variation in findings may be due to differences in antibody detection methods, small sample sizes, use of different time points for different subjects, and varying levels of disease severity. For example, disease severity is a known correlate of antibody levels and persistence (15, 18, 19), and different studies include different proportions of mild or severe disease or do not report disease severity levels. Also, different methods are used to detect antibody titers, and neutralizing titers tend to be low more often than anti-SARS-CoV-2 antibody levels (20, 21). Of course, small sample size is a well-known cause of variable data. In addition to mixed findings regarding antibody persistence, many studies do not evaluate clinical correlates of antibody titers and none have evaluated COVID-19 symptoms as potential correlates. Thus, a standardized approach to evaluating anti-SARS-CoV-2 antibodies with uniform time points, multiple antibody tests, and incorporation of clinical and demographic data, including COVID-19 symptoms, would shed light on the development of antibody-based immunity in COVID-19.

To this end, we broadly evaluated the antibody response against SARS-CoV-2 in a clinically diverse COVID-19 convalescent population at five weeks and three months after symptom resolution using three different types of assays: a novel multiplex assay that evaluates IgG levels against four SARS-CoV-2, one SARS-CoV, and four circulating coronavirus antigens, a SARS-CoV-2 spike protein receptor binding domain (RBD)-angiotensin converting enzyme (ACE)2 binding inhibition assay, and a SARS-CoV-2 neutralization assay. We then correlated antibody titers with a variety of clinical and demographic factors including COVID-19 symptoms. Similar to previous studies, we found that disease severity, older age, and male sex correlate with higher anti-SARS-CoV-2 antibody titers. We also identify fever, abdominal pain, diarrhea and low appetite as symptoms that correlate with higher antibody levels against SARS-CoV-2 and demonstrate persistence of anti-SARS-CoV-2 antibodies three months after symptom resolution.

## RESULTS

We recruited 120 COVID-19 convalescent subjects into the University of Wisconsin (UW) COVID-19 Convalescent Biorepository. Seven subjects were excluded from this study due to erroneous blood collection timing (n=1), receipt of convalescent plasma (n=3), or partial consent (n=3). Two additional subjects were excluded from longitudinal evaluation due to a blood draw >14 days from the 3 month time point. Of the included subjects, blood was collected from 113 at 5 weeks (range 29-48 days, median 36 days, IQR 35-39 days) and from 79 at 3 months (range 85-102 days, median 91 days, IQR 90-93 days) post-symptom resolution. Eighty-one percent of COVID-19 convalescent subjects had a primary care appointment within two years of the first blood draw and/or a hospital admission note that included past medical history and medications. Subjects ranged in age from 19-83 years and had a variety of COVID-19 manifestations (Table 1 and Supplementary Figure 1). As expected (18, 22), hospitalized subjects were more likely to be older and male with more comorbidities such as vascular disease, but were less likely to have asthma (Table 1). Additionally, hospitalized subjects were more likely to have fever and were less likely to have chest tightness, sore throat, or headache than non-hospitalized patients. There was no detectable correlation between race, ethnicity, area of deprivation index (ADI), body mass index (BMI), cancer, immunosuppressing medications, or other COVID-19 symptoms and hospitalization, potentially due to low sample size.

**Table 1.**
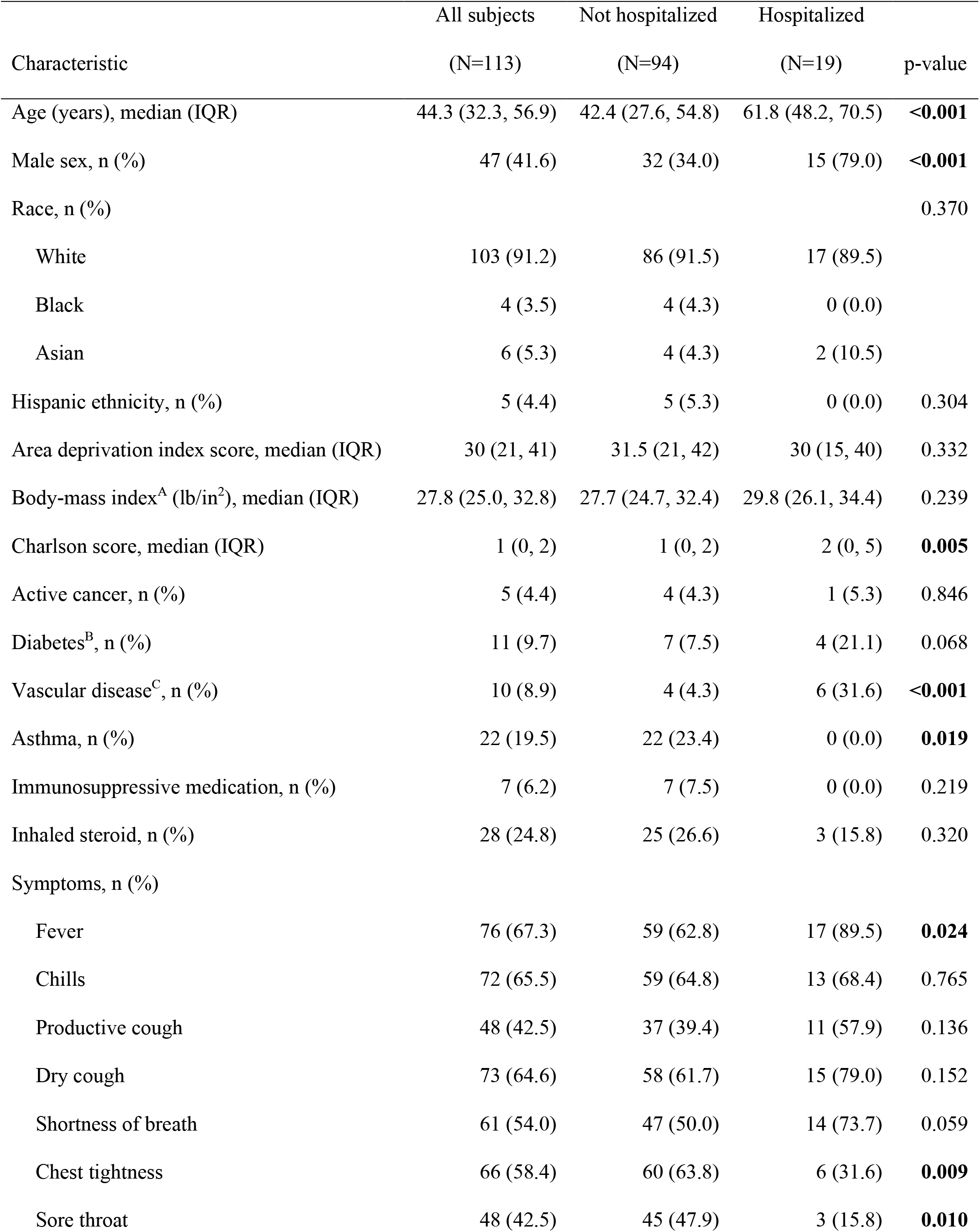
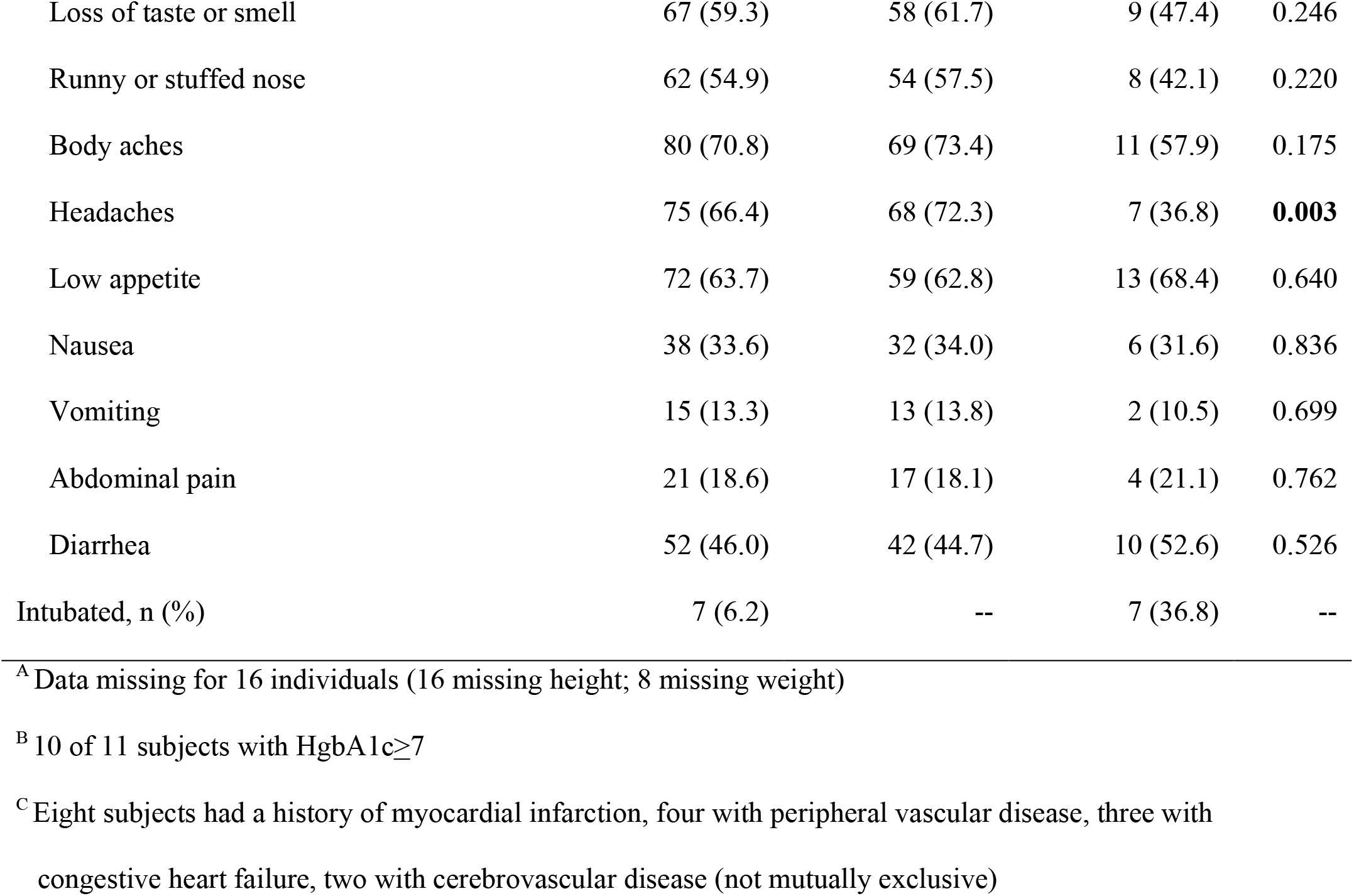
Clinical and demographic characteristics of 113 COVID-19 convalescent subjects.

|  | All subjects | Not hospitalized | Hospitalized |  |
| --- | --- | --- | --- | --- |
| Characteristic | (N=113) | (N=94) | (N=19) | p-value |
| Age (years), median (IQR) | 44.3 (32.3, 56.9) | 42.4 (27.6, 54.8) | 61.8 (48.2, 70.5) | **<0.001** |
| Male sex, n (%) | 47 (41.6) | 32 (34.0) | 15 (79.0) | **<0.001** |
| Race, n (%) |  |  |  | 0.370 |
| White | 103 (91.2) | 86 (91.5) | 17 (89.5) |  |
| Black | 4 (3.5) | 4 (4.3) | 0 (0.0) |  |
| Asian | 6 (5.3) | 4 (4.3) | 2 (10.5) |  |
| Hispanic ethnicity, n (%) | 5 (4.4) | 5 (5.3) | 0 (0.0) | 0.304 |
| Area deprivation index score, median (IQR) | 30 (21, 41) | 31.5 (21, 42) | 30 (15, 40) | 0.332 |
| Body-mass index<sup>A</sup> (lb/in<sup>2</sup>), median (IQR) | 27.8 (25.0, 32.8) | 27.7 (24.7, 32.4) | 29.8 (26.1, 34.4) | 0.239 |
| Charlson score, median (IQR) | 1 (0, 2) | 1 (0, 2) | 2 (0, 5) | **0.005** |
| Active cancer, n (%) | 5 (4.4) | 4 (4.3) | 1 (5.3) | 0.846 |
| Diabetes<sup>B</sup>, n (%) | 11 (9.7) | 7 (7.5) | 4 (21.1) | 0.068 |
| Vascular disease<sup>C</sup>, n (%) | 10 (8.9) | 4 (4.3) | 6 (31.6) | **<0.001** |
| Asthma, n (%) | 22 (19.5) | 22 (23.4) | 0 (0.0) | **0.019** |
| Immunosuppressive medication, n (%) | 7 (6.2) | 7 (7.5) | 0 (0.0) | 0.219 |
| Inhaled steroid, n (%) | 28 (24.8) | 25 (26.6) | 3 (15.8) | 0.320 |
| Symptoms, n (%) |  |  |  |  |
| Fever | 76 (67.3) | 59 (62.8) | 17 (89.5) | **0.024** |
| Chills | 72 (65.5) | 59 (64.8) | 13 (68.4) | 0.765 |
| Productive cough | 48 (42.5) | 37 (39.4) | 11 (57.9) | 0.136 |
| Dry cough | 73 (64.6) | 58 (61.7) | 15 (79.0) | 0.152 |
| Shortness of breath | 61 (54.0) | 47 (50.0) | 14 (73.7) | 0.059 |
| Chest tightness | 66 (58.4) | 60 (63.8) | 6 (31.6) | **0.009** |
| Sore throat | 48 (42.5) | 45 (47.9) | 3 (15.8) | **0.010** |
| Loss of taste or smell | 67 (59.3) | 58 (61.7) | 9 (47.4) | 0.246 |
| Runny or stuffed nose | 62 (54.9) | 54 (57.5) | 8 (42.1) | 0.220 |
| Body aches | 80 (70.8) | 69 (73.4) | 11 (57.9) | 0.175 |
| Headaches | 75 (66.4) | 68 (72.3) | 7 (36.8) | **0.003** |
| Low appetite | 72 (63.7) | 59 (62.8) | 13 (68.4) | 0.640 |
| Nausea | 38 (33.6) | 32 (34.0) | 6 (31.6) | 0.836 |
| Vomiting | 15 (13.3) | 13 (13.8) | 2 (10.5) | 0.699 |
| Abdominal pain | 21 (18.6) | 17 (18.1) | 4 (21.1) | 0.762 |
| Diarrhea | 52 (46.0) | 42 (44.7) | 10 (52.6) | 0.526 |
| Intubated, n (%) | 7 (6.2) | -- | 7 (36.8) | -- |

<sup>A</sup> Data missing for 16 individuals (16 missing height; 8 missing weight)
<sup>B</sup> 10 of 11 subjects with HgbA1c ≥ 7
<sup>C</sup> Eight subjects had a history of myocardial infarction, four with peripheral vascular disease, three with congestive heart failure, two with cerebrovascular disease (not mutually exclusive)

We used a multiplex approach to evaluate IgG levels against SARS-CoV-2 spike (S) protein, RBD of S, N terminal domain (NTD) of S, and nucleocapsid (N), as well as the S protein of SARS-CoV and four seasonal coronaviruses (HCoV-OC43, HCoV-HKU1, HCoV-NL63, HCoV-229E) in subjects five weeks post-COVID-19 symptom resolution. As shown in Figure 1A, COVID-19 convalescent subjects have significantly higher IgG levels against all 4 SARS-CoV-2 antigens compared to SARS-CoV-2 naive subjects. Further, 98% of COVID-19 convalescent subjects had higher binding in at least one test than any naive subject. Finally, IgG levels against S from SARS-CoV and the seasonal betacoronaviruses HCoV-OC43 and HCoV-HKU1, but not the seasonal alphacoronaviruses HCoV-NL63 and HCoV-229E, were higher in COVID-19 convalescent subjects compared to naive controls.

**Figure 1.**
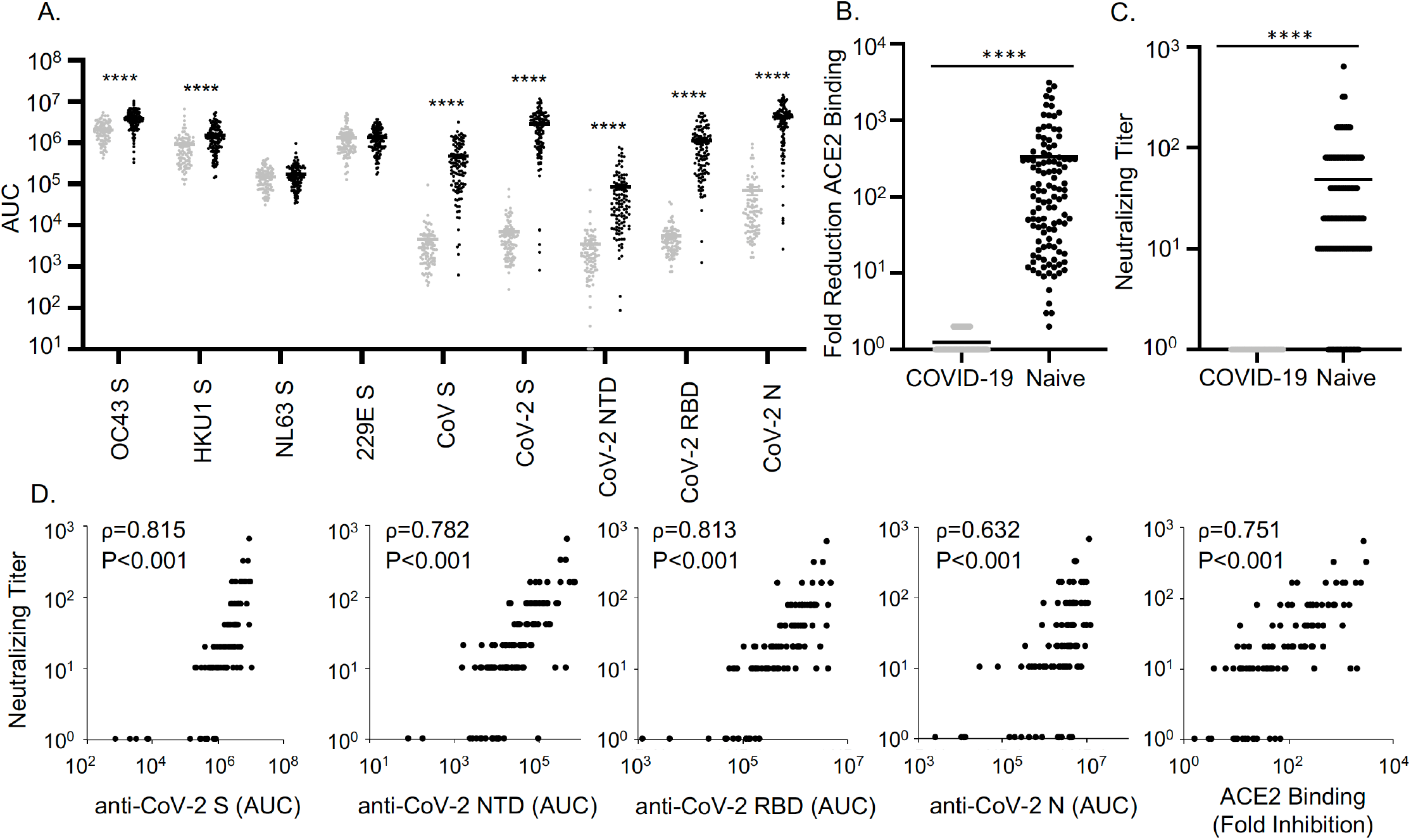
Antibodies against SARS-CoV-2 in COVID-19 convalescent subjects five weeks post-symptom resolution. A. Sera from COVID-19 convalescent (black dots, n=113) and SARS-CoV-2 naive (gray dots, n=87) subjects were analyzed with a multiplex assay to quantify IgG that bound to seasonal HCoV-OC43, HCoV-HKU1, HCoV-NL63, and HCoV-229E spike (S) protein, SARS-CoV S, and SARS-CoV-2 S, N terminal domain (NTD) of S, receptor binding domain (RBD) of S, and nucleocapsid (N) protein. Graph depicts IgG levels reported as area under the curve (AUC). Unpaired t test with Welch’s correction was used to compare COVID-19 convalescent vs. naive means for each antigen. B. Fold reduction of angiotensin converting enzyme (ACE)2 binding to RBD was assessed with a RBD-ACE2 binding inhibition assay for COVID-19 convalescent (n=113) and naive (n=88) sera and a t test with Welch’s correction was used to compare means. C. Neutralizing antibody (Nab) titers were compared by t test with Welch’s correction for COVID-19 convalescent (n=113) and naive (n=30) sera. D. Nab titers were compared with anti-SARS-CoV-2 IgG levels and RBD-ACE2 binding inhibition for COVID-19 convalescent subjects (n=113). Spearman correlation coefficient (ρ) and p value are listed. For all panels, bars represent mean and ****p<0.0001.

We then evaluated the antibody response against SARS-CoV-2 in a more functional manner. Since the RBD of S binds to ACE2, enabling viral entry (23), we quantified the ability of sera to inhibit RBD binding to ACE2. Compared to SARS-CoV-2 naive sera, COVID-19 five week convalescent sera had a much greater ability to inhibit ACE2 binding to RBD (Figure 1B). Further, in a neutralizing assay using live SARS-CoV-2, COVID-19 convalescent sera had higher neutralizing antibody (Nab) titers compared to SARS-CoV-2 naive sera, although 15% of COVID-19 convalescent subjects did not have Nab titers above naive controls (Figure 1C). Overall, Nab titers correlated well with IgG levels against SARS-CoV-2 antigens, as well as with ACE2 binding inhibition (Figure 1D).

Together these data suggest that the vast majority of COVID-19 patients have antibodies five weeks post-symptom resolution that are able to bind to SARS-CoV-2 proteins, inhibit RBD-ACE2 binding, and neutralize SARS-CoV-2. Also, IgG titers, particularly against S and RBD, and RBD-ACE2 binding inhibition better differentiate between COVID-19 convalescent and naive individuals than the neutralizing assay, but may overestimate protective antibodies.

We then evaluated if antibody titers at five weeks post-symptom resolution varied with disease severity in our cohort. As shown in Figure 2, hospitalized subjects had higher antibody levels than non-hospitalized subjects according to all of our tests. Moreover, in general, antibody titers increased with COVID-19 severity (Figure 2). We next evaluated if clinical and demographic factors apart from disease severity correlated with antibody titers. Since multiple variables correlate with hospitalization (Table 1), a marker of severe disease, we analyzed non-hospitalized subjects only. As shown in Table 2, older age, male sex, higher BMI and a Charlson Comorbidity Index score >2 correlated with higher antibody titers in non-hospitalized subjects for all or most tests. However, race, ethnicity, ADI, active cancer, diabetes mellitus, vascular disease, asthma, immunosuppressive medications and inhaled/intranasal steroids did not correlate with antibody titers in general (Supplementary Table 1). Also, in non-hospitalized subjects, fever, low appetite, abdominal pain, and diarrhea correlated with higher antibody levels measured by every test, whereas cough, body aches, headache, nausea, and vomiting only correlated with some antibody tests and chills, shortness of breath, chest tightness, sore throat, loss of taste or smell, and runny or stuffed nose correlated with no or almost no antibody tests (Table 3). Together, these data suggest that several demographic and clinical factors, including COVID-19 symptoms, correlate with increased antibody titers against SARS-CoV-2.

**Table 2.** Median (IQR) IgG antibody levels, fold RBD-ACE2 inhibition, and SARS-CoV-2 neutralization titers five weeks after resolution of COVID-19 symptoms according to clinical and demographic characteristics in non-hospitalized subjects.

|  | N | Anti-Spike <sup>A</sup> | Anti-NTD <sup>A</sup> | Anti-RBD <sup>A</sup> | Anti-N <sup>A</sup> | ACE-2 Inhib. | Nab Titer |
| --- | --- | --- | --- | --- | --- | --- | --- |
| All subjects | 94 | 19.3 (7.0, 35.2) | 0.27 (0.10, 0.69) | 5.1 (1.8, 12.3) | 31.3 (11.0, 51.1) | 72.5 (20, 287) | 20 (10, 40) |
| Age quartile |  | **p=0.047** | **p=0.009** | **p=0.028** | **p=0.005** | **p=0.011** | p=0.230 |
| 1 (19.2-27.6) | 24 | 10.0 (3.8, 22.4) | 0.14 (0.07, 0.42) | 2.2 (1.2, 7.3) | 9.6 (3.2, 34.7) | 32 (10.5, 94) | 10 (10, 20) |
| 2 (29.6-42.4) | 23 | 17.8 (8.4, 28.3) | 0.25 (0.15, 0.53) | 5.3 (1.8, 9.6) | 29.0 (22.8, 55.8) | 85 (50, 203) | 20 (10, 40) |
| 3 (42.5-54.5) | 23 | 31.0 (7.9, 36.5) | 0.51 (0.08, 1.0) | 10.0 (2.1, 14.9) | 37.4 (18.8, 51.1) | 119 (15, 311) | 20 (1, 40) |
| 4 (54.8-76.5) | 24 | 29.9 (8.8, 46.3) | 0.37 (0.20, 1.7) | 8.4 (3.5, 22.4) | 44.1 (28.5, 62.7) | 216 (35.5, 555) | 20 (10, 80) |
| Sex |  | **p=0.034** | **p=0.039** | **p=0.015** | **p=0.031** | **p=0.023** | **p=0.009** |
| Male | 32 | 29.4 (7.2, 43.0) | 0.46 (0.15, 0.89) | 9.6 (2.5, 16.3) | 44.4 (23.0, 65.9) | 121 (42.5, 343.5) | 40 (10, 80) |
| Female | 62 | 16.8 (7.0, 32.0) | 0.21 (0.08, 0.47) | 4.1 (1.6, 10.0) | 29.2 (8.7, 44.4) | 50.5 (15, 219) | 15 (10, 40) |
| BMI <sup>B</sup> (lb/in<sup>2</sup>) |  | **p=0.041** | **p=0.011** | **p=0.020** | p=0.091 | p=0.067 | **p=0.027** |
| 1 (<25) | 22 | 10.9 (5.0, 20.1) | 0.16 (0.09, 0.27) | 3.1 (1.3, 6.4) | 22.4 (10.4, 39.9) | 38 (20, 121) | 10 (10, 20) |
| 2 (25-29.99) | 27 | 20.9 (7.9, 43.1) | 0.41 (0.12, 1.02) | 6.9 (2.2, 15.8) | 42.9 (16.1, 51.1) | 72 (17, 328) | 10 (10, 40) |
| 3 (≥30) | 30 | 27.4 (15.0, 38.3) | 0.41 (0.18, 0.86) | 9.1 (4.8, 13.4) | 43.5 (23.3, 85.1) | 138.5 (45, 298) | 40 (10, 80) |
| CCIS <sup>C</sup> |  | **p=0.021** | **p=0.001** | **p=0.010** | **p=0.010** | **p=0.009** | p=0.072 |
| > 2 | 30 | 33.2 (8.8, 45.1) | 0.46 (0.25, 1.4) | 9.8 (4.1, 20.2) | 45.5 (30.4, 62.8) | 216.5 (34, 489) | 30 (10, 80) |

<sup>A</sup> Values are x10<sup>5</sup>
<sup>B</sup> Data missing for 15 subjects
<sup>C</sup> Charlson Comorbidity Index score

**Table 3.** Median (IQR) IgG antibody levels, fold RBD-ACE2 inhibition, and SARS-CoV-2 neutralization titers five weeks after resolution of COVID-19 symptoms according to clinical symptoms in non-hospitalized subjects.

|  | N | Anti-Spike <sup>A</sup> | Anti-NTD <sup>A</sup> | Anti-RBD <sup>A</sup> | Anti-N <sup>A</sup> | ACE-2 Inhib. | Nab Titer |
| --- | --- | --- | --- | --- | --- | --- | --- |
| All subjects | 94 | 19.3 (7.0, 35.2) | 0.27 (0.10, 0.69) | 5.1 (1.8, 12.3) | 31.3 (11.0, 51.1) | 72.5 (20, 287) | 20 (10, 40) |
| Fever | 59 | **28.6 (7.9, 39.1)\*\*** | **0.41 (0.17, 0.86)\*\*** | **9.2 (2.8, 14.2)\*\*** | **39.7 (22.8, 54.4)\*** | **121 (37, 328)\*\*** | **20 (10, 80)\*\*\*** |
| Chills <sup>B</sup> | 59 | 20.9 (7.9, 38.7) | 0.32 (0.14, 0.82) | 5.9 (2.1, 14.0) | 38.3 (16.1, 52.9) | 73 (34, 305) | **20 (10, 80)\*** |
| Productive cough | 37 | 26.3 (10.5, 38.0) | **0.44 (0.18, 0.74)\*** | **8.7 (3.2, 13.4)\*** | 41.8 (22.8, 55.2) | **131 (42, 305)\*** | **20 (10, 80)\*\*** |
| Dry cough | 58 | **26.4 (10.3, 38.0)\*\*** | **0.32 (0.17, 0.73)\*** | **7.4 (2.8, 12.9)\*\*** | 38.6 (21.4, 52.8) | **124 (37, 298)\*** | **20 (10, 40)\*** |
| Shortness of breath | 47 | 26.3 (6.7, 38.3) | 0.27 (0.12, 0.74) | 5.9 (1.8, 12.8) | 26.7 (11.0, 55.8) | 85 (22, 305) | 20 (10, 80) |
| Chest tightness | 60 | 20.7 (7.2, 35.4) | 0.26 (0.09, 0.71) | 5.8 (1.8, 12.5) | 35.6 (11.7, 55.5) | 79 (21, 287) | 20 (10, 40) |
| Sore throat | 45 | 22.4 (8.4, 35.2) | 0.34 (0.17, 0.68) | 6.9 (2.4, 11.8) | 38.4 (14.2, 52.8) | 73 (20, 298) | 20 (10, 40) |
| Loss of taste or smell | 58 | 20.7 (8.0, 35.6) | 0.33 (0.10, 0.73) | 5.0 (1.8, 12.9) | 30.6 (13.9, 54.4) | 72.5 (22, 298) | 20 (10, 40) |
| Runny or stuffed nose | 54 | 20.0 (6.7, 36.5) | 0.28 (0.09, 0.69) | 5.5 (1.7, 11.8) | 34.8 (16.1, 52.9) | 72.5 (20, 279) | 20 (10, 40) |
| Body aches | 69 | **24.5 (8.4, 38.7)\*\*** | **0.29 (0.14, 0.86)\*** | **6.9 (2.2, 14.0)\*\*** | 38.4 (16.1, 52.9) | **93 (28, 298)\*** | **20 (10, 80)\*** |
| Headaches | 68 | **24.3 (9.1, 35.4)\*** | **0.31 (0.16, 0.71)\*** | **6.8 (2.9, 12.9)\*\*** | 36.2 (18.4, 52.8) | 91.5 (26, 292.5) | **20 (10, 40)\*** |
| Low appetite | 59 | **26.5 (9.8, 43.1)\*\*\*** | **0.41 (0.18, 1.02)\*\*\*** | **8.7 (2.9, 15.8)\*\*\*** | **39.7 (22.2, 55.8)\*\*** | **123 (28, 311)\*\*** | **20 (10, 80)\*\*\*** |
| Nausea | 32 | **28.7 (9.2, 45.5)\*** | **0.45 (0.26, 1.07)\*\*** | **9.1 (3.5, 14.2)\*** | **42.3 (25.8, 56.1)\*** | 95 (18.5, 331.5) | 20 (10, 80) |
| Vomiting | 13 | **30.2 (19.5, 43.3)\*** | **0.93 (0.44, 1.14)\*\*\*** | **10.5 (4.8, 20.2)\*** | **48.2 (41.8, 82.2)\*\*** | 131 (26, 376) | 40 (10, 80) |
| Abdominal pain | 17 | **34.9 (16.9, 46.5)\*** | **1.02 (0.28, 1.64)\*\*** | **14.0 (5.6, 20.2)\*** | **48.2 (28.8, 69.4)\*** | **310 (47, 756)\*** | **80 (20, 80)\*\*** |
| Diarrhea | 42 | **28.4 (13.2, 42.8)\*\*** | **0.44 (0.23, 0.93)\*\*** | **8.9 (4.1, 14.2)\*** | **40.6 (23.3, 82.2)\*\*** | **124 (47, 392)\*\*** | **30 (10, 80)\*** |

<sup>A</sup> Values are x10<sup>5</sup>
<sup>B</sup> Data missing for three subjects
\*p<0.05, \*\*p<0.01, \*\*\*p<0.001

**Figure 2.**
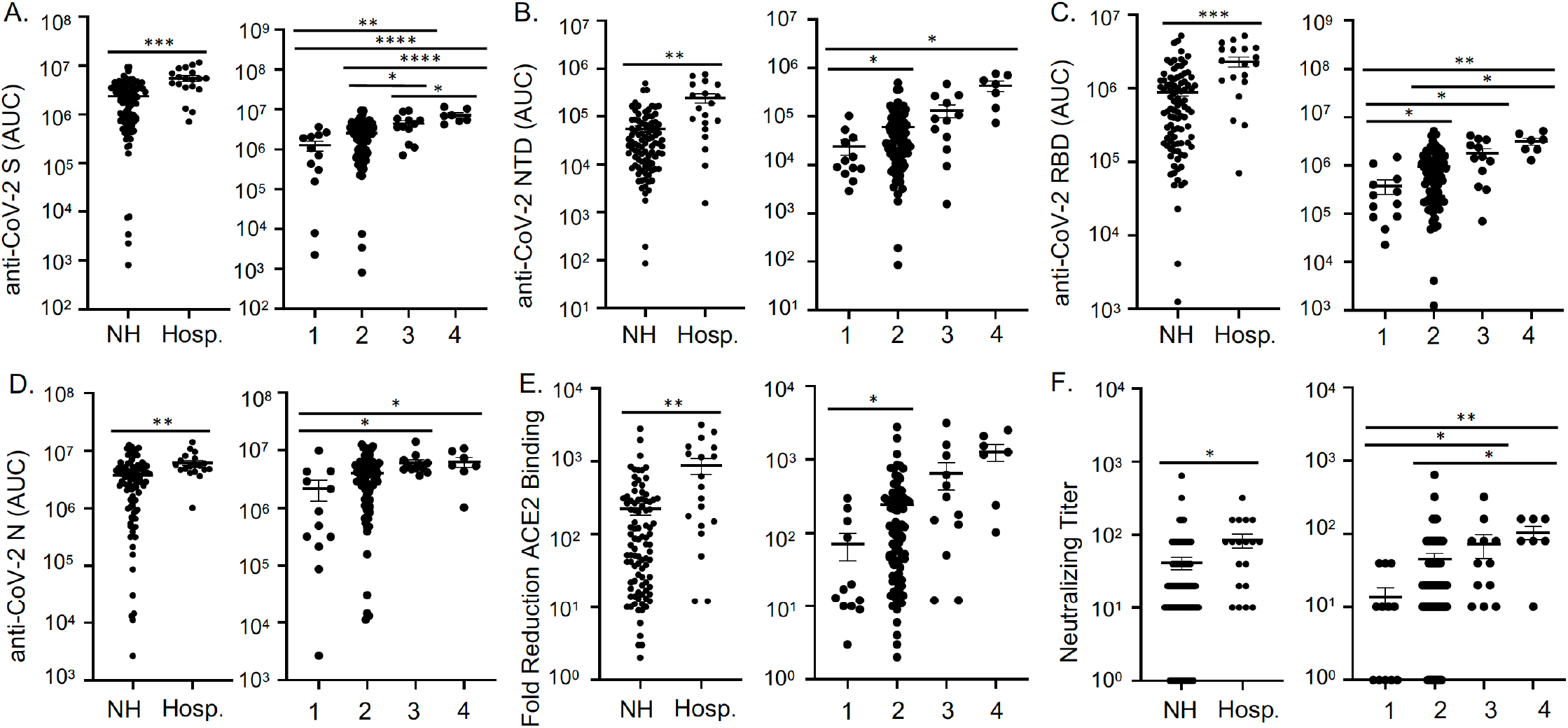
Patients with more severe COVID-19 have higher antibody levels against SARS-CoV-2. IgG levels against SARS-CoV-2 S (A), NTD (B), RBD (C), and N (D) determined by multiplex assay, as well as fold reduction of RBD-ACE2 binding (E), and neutralizing antibodies (F) in COVID-19 convalescent sera five weeks post-symptom resolution were compared for non-hospitalized (NH) versus hospitalized (Hosp.) subjects by t test and compared among subjects with differing COVID-19 severity scores by ANOVA. Anti-NTD, anti-RBD, and fold reduction of RBD-ACE2 binding were compared by Welch’s ANOVA with Dunnett’s test. Anti-S and anti-N were analyzed by ANOVA with Tukey’s multiple comparisons test, and neutralizing titers were analyzed by Kruskal-Wallis ANOVA with Dunn’s multiple comparisons test. For all panels: n=19 Hosp., 94 NH, 12 with severity score 1 (no fever, chills, productive cough, shortness of breath, or hospitalization), 82 with score 2 (fever, chills, productive cough, or shortness of breath, but no hospitalization), 12 with score 3 (hospitalized, but not intubated), 7 with score 4 (intubated); lines indicate mean +/- SEM; *p<0.05, **p<0.01, ***p<0.001, ****p<0.0001.

Finally, we evaluated the antibody response against SARS-CoV-2 and other coronaviruses in our COVID-19 convalescent subjects at three months post symptom resolution. We found that in our cohort as a whole, IgG levels against SARS-CoV-2 S, NTD, RBD or N did not decline, nor did RBD-ACE2 binding inhibition or Nab titers from five weeks to three months post-symptom resolution (Figure 3). Titers against SARS-CoV and seasonal coronaviruses also did not fall (Supplementary Figure 2). However, when hospitalized and non-hospitalized subjects were analyzed separately (Figure 3), IgG levels against S and N rose slightly in hospitalized subjects (area under the curve: 4,525,184 +/- 796,452 versus 5,247,312 +/- 811,346 and 5,765,064 +/- 588,170 versus 7,004,295 +/- 654,857, respectively) and Nab titers decreased slightly in non-hospitalized subjects (titers: 49 +/- 11 versus 37 +/-10) over time. Together these data suggest that, in general, antibodies against SARS-CoV-2 persist at least three months after symptom resolution, particularly in the most severely ill patients.

**Figure 3.**
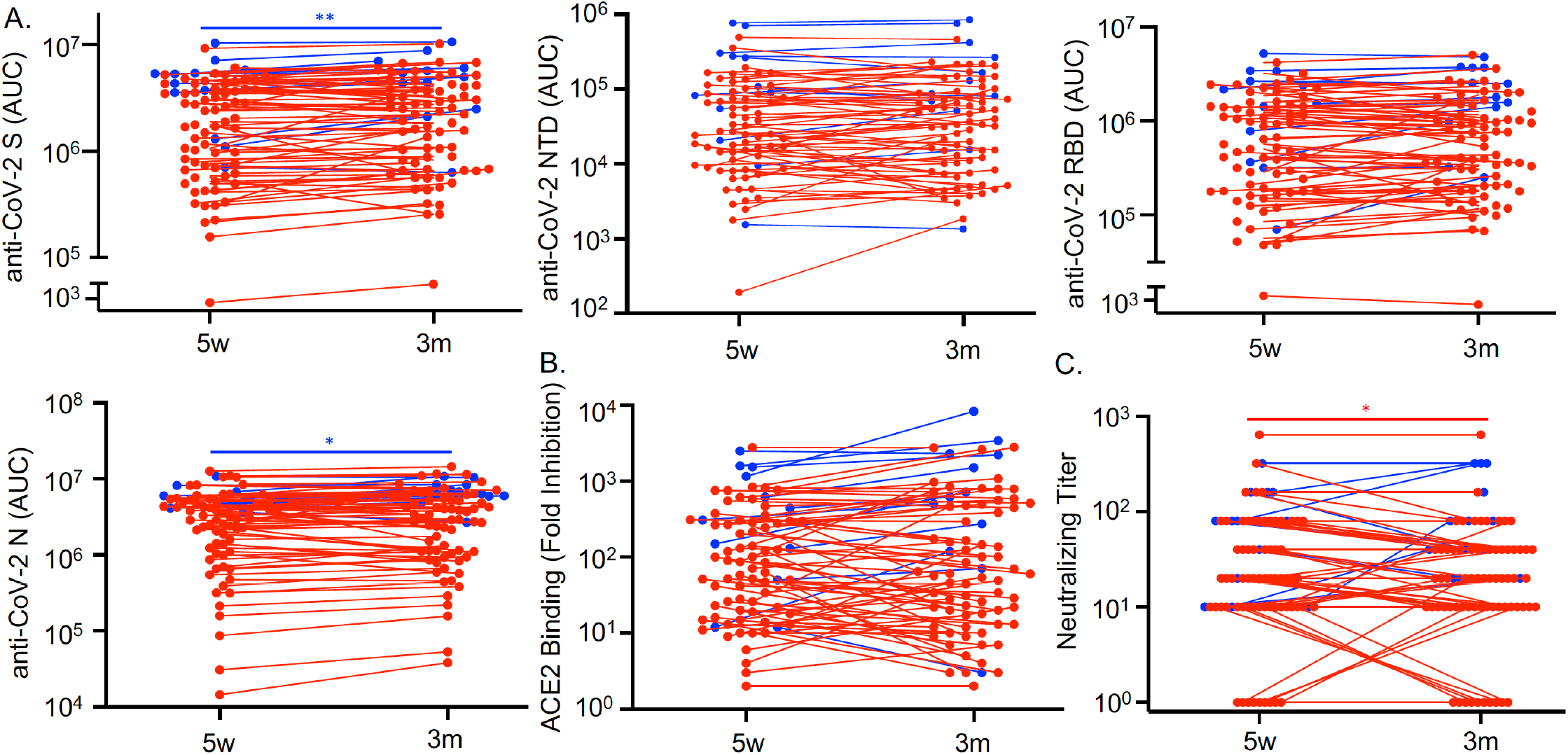
Antibodies against SARS-CoV-2 persist three months after COVID-19 symptom resolution. Sera from COVID-19 convalescent subjects (n=79) collected 5 weeks (w) and 3 months (m) after symptom resolution were subjected to multiplex assay to detect IgG that binds to SARS-CoV-2 S, NTD, RBD and N antigens (A), to RBD-ACE2 binding inhibition assay (B), and to SARS-CoV-2 neutralization assay (C). Dots, lines, and asterisks in red represent non-hospitalized (n=67) and in blue represent hospitalized (n=12) subjects with lines connecting the two time points for individual subjects (*p<0.05 and **p<0.01 by paired t test).

## DISCUSSION

We have demonstrated that the vast majority of COVID-19 convalescent subjects generate antibodies against SARS-CoV-2 that can inhibit ACE2 binding, neutralize live SARS-CoV-2, and persist at least three months post-COVID-19 symptom resolution. Further, we demonstrated that greater disease severity, older age, male sex, higher BMI, higher Charlson Comorbidity Index score, fever, abdominal pain, diarrhea and low appetite all correlate with higher antibody titers using a variety of tests.

Although we detected IgG against SARS-CoV-2 in the majority of COVID-19 convalescent subjects, there was some variability among tests (Figure 1). Using a multiplex assay, IgG levels against S and RBD, but not NTD or N, showed an impressive difference in titers between COVID-19 convalescent and control sera. Similar strong results were observed for the RBD-ACE2 binding inhibition assay. Thus, these three tests could be developed into a diagnostic test with very high sensitivity and specificity. Of note, two COVID-19 convalescent subjects had similar values to naive subjects in all three tests. It is unknown if these subjects had false positive SARS-CoV-2 PCR tests, if they did not make antibodies, or if our tests were insufficiently sensitive to detect their antibodies. Further studies are needed with more asymptomatic subjects and larger sample sizes to better evaluate these tests.

All of the antibody tests correlated well with Nab titers (Figure 1), a gold standard for protective antibodies. This correlation is encouraging, suggesting that our other antibody assays are measuring relevant antibodies without the cost, hazards, time, and expertise needed for neutralizing assays. However, similar to related studies (20, 21), more COVID-19 convalescent subjects had antibody titers no higher than naive controls using the neutralizing assay as compared to other assays. It is unknown if these subjects truly lack protective antibodies or if the neutralization assay is insufficiently sensitive. Future studies are needed to standardize neutralizing assays and to determine truly protective levels of Nab titers.

In addition to the development of anti-SARS-CoV-2 antibodies in COVID-19 convalescent sera five weeks post-symptom resolution, we saw a small increase in antibodies that bind to the spike protein of seasonal betacoronaviruses (OC43 and HKU1), but not seasonal alphacoronaviruses (NL63 and 229E) (Figure 1), perhaps because SARS-CoV-2 is a betacoronavirus (23). Moreover, antibodies that bind to the spike protein of all of the seasonal coronaviruses appeared to slightly rise from five weeks to three months post-symptom resolution (Supplementary Figure 2). A similar “back boost” phenomenon has been reported for other viruses (24) and could represent cross-reactivity of newly developed anti-SARS-CoV-2 antibodies or stimulation of memory B cells that originally developed in response to circulating coronaviruses. In contrast, the higher titers against SARS-CoV S seen in COVID-19 convalescent subjects (Figure 1), which did not change over time (Supplementary Figure 2), are probably due to cross-reactive anti-SARS-CoV-2 antibodies, since it is extremely unlikely that the Wisconsin cohort was exposed to SARS-CoV. The clinical implications of these phenomena are unclear and warrant further investigation.

Similar to recent reports (18, 19, 25-27), we demonstrated that anti-SARS-CoV-2 antibodies were highest in patients with the highest disease severity (Figure 2). We also found that in non-hospitalized patients, higher antibody levels correlated with older age, male sex, higher BMI, and higher Charlson Comorbidity Index score (Table 2). All of these factors correlated with severe disease in our cohort as measured by hospitalization (Table 1) except for BMI, which has been reported by others to be associated with severe disease (28). Thus, non-hospitalized subjects with these high-risk characteristics might have had relatively severe disease that our methods could not measure, driving the higher antibody titers. Alternatively, these characteristics could contribute specifically to increased antibody levels. However, both older age and obesity appear to impair antibody responses (29), and there is no broadly generalizable increased antibody response in males (30). Of note, the correlation between anti-SARS-CoV-2 antibody titers with age, sex, and obesity has been previously reported (19, 31), but we are the first to report a correlation with the Charlson Comorbidity Index score.

In addition to disease severity, we report for the first time that specific COVID-19 symptoms, namely fever, low appetite, abdominal pain, and diarrhea, consistently correlate with higher antibody titers (Table 3). Other symptoms did not correlate consistently or at all including shortness of breath, which a previous study of 68 COVID-19 convalescent subjects who donated plasma at various times post-infection found was associated with higher antibody titers (20). Fever and low appetite are signs of a systemic inflammatory response, suggesting that such an inflammatory response may be key for developing a strong anti-SARS-CoV-2 antibody response. The link between diarrhea and abdominal pain and the antibody response is intriguing. SARS-CoV-2-induced diarrhea may directly exacerbate inflammation and disease severity (32). Thus, diarrhea simply may be a marker of severe disease, which is associated with higher antibody titers. However, we did not see increased diarrhea in our hospitalized patients (Table 1). Another possibility is that diarrhea, which is typically caused by enteric viral infection and damage (32), may directly enhance the antibody response, perhaps by activating inflammatory cells throughout the gut. However, at this time, the mechanism by which diarrhea might enhance antibody titers is unknown and requires further investigation.

Finally, we found that antibodies persist from five weeks until three months after symptom resolution (Figure 3). Some antibody levels even continued to rise slightly during this time period, although Nab titers fell slightly in non-hospitalized subjects. Our findings are discrepant from some studies, which reported falling IgG titers against SARS-CoV-2 within a few months of symptom onset (11-17). However, these studies all had smaller sample sizes, particularly at later time points, or the fall in titers was observed in subjects with mild or asymptomatic disease. Very recently, a small drop in anti-SARS-CoV-2 antibody titers in 121 COVID-19 convalescent subjects from ∼30 to ∼148 days post-symptom onset was reported (33). It is hard to compare our observations with theirs, since clinical characteristics of their subjects were not described. However, our findings are consistent with preliminary reports of persistent antibody titers at least 6 months after disease (8-10). Further studies are needed at later time points, combined with clinical correlates and data about re-infection, in order to assess long-term protective antibody-based immunity against SARS-CoV-2.

There are a few caveats to our studies. First, COVID-19 symptoms were self-reported up to a month after symptom resolution, which could lead to recall error. Additionally, some of the medical records were incomplete with no recent primary care or admission note in the charts of 19% of subjects and 14% missing BMI data. This gap would be biased toward non-hospitalized patients. Also, our population was relatively homogeneous with regard to race and ethnicity. However, our study is strong in that it includes a wide breadth of COVID-19 severity in 113 subjects with consistent time points and multiple types of antibody tests.

In sum, we report that antibody-based immunity against SARS-CoV-2 lasts at least three months post-symptom resolution and that antibody titers consistently correlate with specific COVID-19 symptoms (fever, low appetite, abdominal pain, and diarrhea) and clinical and demographic features (older age, male sex, higher COVID-19 severity, higher BMI, and higher Charlson Comorbidity Index score). Future work is needed to determine which antibody titers are protective against re-infection and how long those titers last.

## METHODS

### Human Subjects

COVID-19 convalescent sera and data were obtained from the UW COVID-19 Convalescent Biorepository and control sera collected prior to 2019 were obtained from the UW Rheumatology Biorepository (34), which includes subjects with and without rheumatologic disease, and from the NIH clinical protocol VRC200. All subjects provided informed consent. For the COVID-19 Convalescent Biorepository, all individuals 18+ years old who tested positive for SARS-CoV-2 by PCR at UW Health were invited to participate until 120 subjects were recruited. Clinical and demographic data were collected by survey upon recruitment. Additional data and blood were collected 5 weeks and 3 months +/- 10 days post-symptom resolution. Age, sex, address (to determine ADI (35)), medications, lab values, height and weight (to calculate BMI), medical problems, and date of most recent primary care appointment were abstracted from the UW Health electronic medical record (EMR). Race, ethnicity, tobacco use, COVID-19 symptoms, and date of symptom resolution were self-reported by questionnaire. Hospitalization and intubation status for COVID-19 were obtained by questionnaire and EMR abstraction. COVID-19 severity was scored as critical (4, intubated), severe (3, hospitalized but not intubated), moderate (2, fever defined as temperature >100°F, chills, productive cough, or shortness of breath, but not hospitalized), or mild (1, none of the above). Charlson Comorbidity Index Scores were calculated (36). Subjects were excluded from this study if they received convalescent plasma, if blood was collected more than 14 days from the intended time point, or if they did not provide consent for all aspects of the study.

### IgG binding to coronavirus antigens (multiplex assay)

Plates (96 well) printed with SARS-CoV-2 S protein, RBD of S, NTD of S, and N protein as well as S from SARS-CoV, HCoV-HKU1, HCoV-OC43, HCoV-NL63, and HCoV-229E, in addition to bovine serum albumin (BSA) were supplied by Meso Scale Discovery (MSD, Rockville, USA). Plates were blocked for 60 minutes with MSD Blocker A (5% BSA) followed by washing. Then, sera were applied to the wells at 4 dilutions (1:100, 1:800, 1:3200 and 1:12,800) and incubated with shaking for 2 hours. Plates were washed and SULFO-TAG labeled anti-IgG (MSD) was applied to the wells for 1 hour. Plates were washed, enhanced chemiluminescence (ECL) substrate (MSD) applied, and light emission (as a measure of bound IgG) read with the MSD Sector instrument. BSA readings were subtracted from CoV antigen readings. Area under the curve values for each sample were used for statistical analysis with zero values (3 samples for anti-NTD) depicted as ten in graphs for optimal visualization using the log scale.

### RBD-ACE2 binding inhibition assay

Plates (384 well) precoated with RBD were supplied by MSD. Plates were blocked for 30 minutes with MSD Blocker A, washed, sera applied at 1:10 dilution, and incubated with shaking for 1 hour. SULFO-TAG labeled ACE2 was applied to the wells, incubated for 1 hour, and washed. ECL substrate was applied, and light emission (a measure of RBD-ACE2 complex) was read by the MSD Sector instrument. The amount of light emitted in wells containing no sample (assay diluent only) was considered the maximal binding response. Reduction of ECL response from the maximal binding response was directly proportional to the extent of competitive binding activity.

### Neutralization assay

Vero E6/TMPRSS2 cells were grown in Dulbecco’s Modified Eagle Medium supplemented with 5% fetal calf serum, HEPES, amphotericin B, and gentamicin sulfate. Sera (100µl) were diluted in cell culture solution with 2-fold serial dilutions from 5x to 2560x. Virus (SARS-CoV-2/UW-001/Human/2020/Wisconsin) was diluted in cell culture solution to an adjusted titer of 100 plaque-forming units (PFU) per 60µl. Diluted sera (60µl) and diluted virus (60µl) were mixed in wells of 96-well U-bottom plates in duplicate. Plates were incubated at 37°C for 30 minutes. Culture supernatant was aspirated from Vero E6/TMPRSS2 cells plated in 96 well dishes and replaced with the mixtures of serially diluted sera and virus (100µl/well, in duplicate) followed by incubation at 37°C with 5% CO2 for 3 days. Crystal violet stain was added to wells to stain for living cells. Neutralization titers were determined by the maximum fold dilution at which the serum samples could completely prevent cell death as determined by eye. Some duplicates diverged by a single fold dilution in their neutralization titer. In this situation, the lower dilution was used as the neutralization titer. Sera with cell death at all dilutions were assigned a dilution value of 1 for analysis purposes.

### Statistics

Antibody levels were compared between COVID-19 convalescent and control sera using a t test or among subsets of COVID-19 convalescent sera by one-way ANOVA with Tukey’s, Dunnett’s, or Dunn’s multiple comparisons tests as appropriate. Welch’s correction was used for all tests when there was unequal variance. Serum anti-SARS-CoV-2 antibody levels at five weeks and three months post symptom resolution in the same subjects were compared using paired t tests. Correlations between antibody titers obtained by different tests were estimated by Spearman rank correlation coefficients. The relationship between clinical and demographic factors (age, sex, race, ethnicity, ADI, BMI, Charlson Comorbidity Index score, active cancer, diabetes, vascular disease, asthma, immunosuppressive medications, inhaled/intranasal steroid, symptoms, and intubation status) and having been hospitalized for COVID-19 were examined using the Pearson’s chi-squared test for categorical variables and Kruskal-Wallis test for non-normally distributed continuous data. Similar analyses were performed in non-hospitalized COVID-19 recovered individuals alone to correlate antibody levels with clinical and demographic features. Analyses were performed using GraphPad Prism software (San Diego, USA) and STATA version 16 (College Station, USA). For all analyses, p<0.05 was considered significant.

### Study approval

Human studies were performed according to the Declaration of Helsinki and were approved by the Institutional Review Board at the UW-Madison. Written informed consent was received from participants prior to inclusion in the study.

## Data Availability

Data will be provided upon reasonable request.

## AUTHOR CONTRIBUTIONS

MAS conceived of and oversaw the study. MFA, AMM, SJB and CRG prepared COVID-19 convalescent sera. MFA and ASH abstracted information from the EMR. SEO, SRN, AT, and BF developed and performed multiplex and inhibition assays under the guidance of ABM. TA and PJH developed and performed neutralizing assays under the guidance of YK. MFA and AKS performed statistical analyses. MFA and MAS wrote the manuscript which was reviewed, edited, and approved by all authors.

## ACKNOWLEDGEMENTS

This project was funded through a COVID-19 Response grant from the Wisconsin Partnership Program and a pilot award from the Department of Medicine both at the University of Wisconsin School of Medicine and Public Health to M.A.S. Additional support was provided by the National Institutes of Health National Institute on Aging to M.F.A. (T32 AG000213), National Heart, Lung, and Blood Institute (T32 HL007899) to A.M.M., and National Institute of Allergy and Infectious Disease to A.S.H. (T32 AI007414).

