## Supplementary for "Fever, Diarrhea, and Severe Disease Correlate with High Persistent Antibody Levels against SARS-CoV-2"

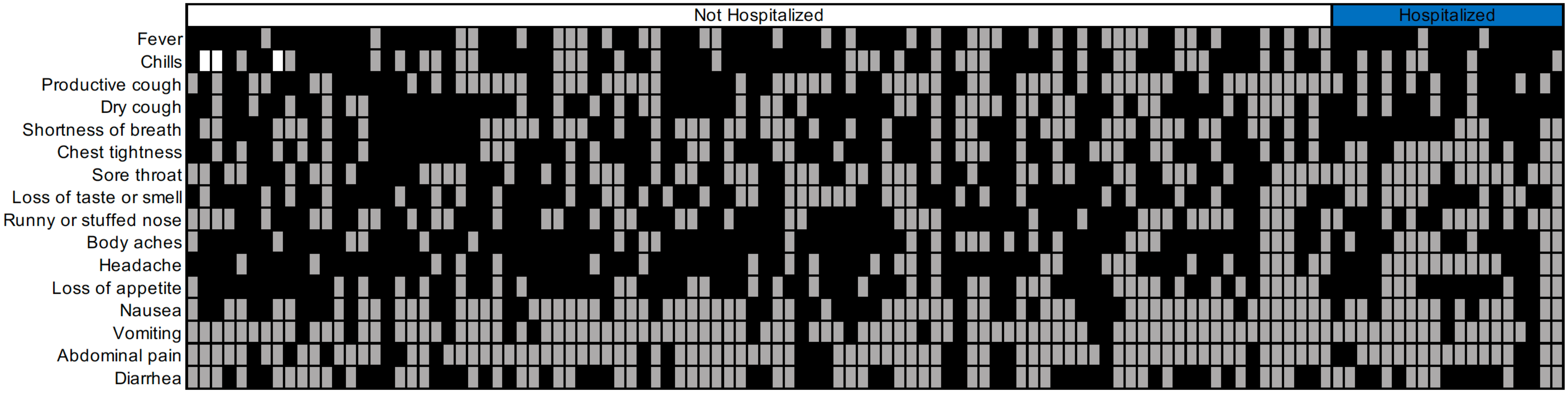


**Supplementary Figure 1. Self-reported symptoms during COVID-19.** Upon initial recruitment, subjects were asked about each symptom listed on the left during their COVID-19 illness. Each column represents a subject (n=113) with hospitalized subjects under the blue bar. Black square, symptom experienced; gray square, symptom not experienced; white square, data not collected.

**Supplemental Table 1. Median (IQR) IgG antibody levels, fold RBD-ACE2 inhibition, and SARS-CoV-2 neutralization titers five weeks after resolution of COVID-19 symptoms according to clinical and demographic characteristics in non-hospitalized subjects**

| | N | Anti-S [A] | Anti-NTD [A] | Anti-RBD [A] | Anti-N [A] | ACE-2 inhib. | Nab Titer |
|---|---|---|---|---|---|---|---|
| All subjects | 94 | 19.3 (7.0, 35.2) | 0.27 (0.10, 0.69) | 5.1 (1.8, 12.3) | 31.3 (11.0, 51.1) | 72.5 (20, 287) | 20 (10, 40) |
| Race | | p=0.580 | p=0.716 | p=0.530 | p=0.179 | p=0.683 | p=0.235 |
| White | 86 | 18.4 (6.7, 34.9) | 0.25 (0.09, 0.73) | 4.9 (1.7,11.8) | 30.0 (10.4, 51.1) | 71.5 (22, 268) | 20 (10, 40) |
| Black | 4 | 26.1 (14.3, 39.9) | 0.31 (0.23, 0.86) | 7.0 (3.4, 14.3) | 51.7 (40.6, 86.2) | 186 (15.5, 421) | 30 (15, 60) |
| Asian | 4 | 31.2 (15.2, 40.5) | 0.47 (0.20, 0.67) | 10.4 (4.2, 19.2) | 25.8 (14.6, 34.2) | 345.5 (75, 568.5) | 60 (25, 120) |
| Hispanic ethnicity | | **p=0.048** | p=0.097 | p=0.100 | p=0.136 | p=0.056 | p=0.057 |
| | 5 | 39.1 (30.2, 46.0) | 0.77 (0.56, 1.1) | 13.4 (10.5, 14.0) | 52.8 (43.9, 55.2) | 310 (245, 376) | 40 (40, 80) |
| ADI Index Score tertile | | p=0.478 | p=0.642 | p=0.471 | p=0.544 | p=0.573 | p=0.662 |
| 1 (4, 24) | 33 | 19.0 (8.4, 31.0) | 0.23 (0.11, 0.66) | 4.8 (1.7, 10.9) | 26.7 (12.4, 50.8) | 52 (17, 131) | 20 (10, 80) |
| 2 (26, 40) | 32 | 15.4 (5.3, 36.0) | 0.26 (0.08, 0.62) | 4.4 (1.6, 12.6) | 30.9 (8.5, 49.4) | 86 (21, 272.5) | 20 (10, 40) |
| 3 (41, 99) | 29 | 22.7 (8.0, 43.3) | 0.32 (0.13, 0.93) | 7.2 (2.8, 14.0) | 42.3 (24.6, 54.4) | 100 (37, 310) | 20 (10, 40) |
| Active cancer | | p=0.190 | p=0.925 | p=0.349 | p=0.779 | p=0.210 | p=0.312 |
| | 4 | 10.3 (3.7, 18.9) | 0.30 (0.18, 0.40) | 3.5 (1.9, 5.4) | 36.0 (18.6, 41.0) | 28 (15.5, 87.5) | 10 (5.5, 25) |
| Diabetes mellitus | | p=0.424 | p=0.589 | p=0.416 | p=0.069 | p=0.074 | p=0.320 |
| | 7 | 33.2 (22.6, 77.3) | 0.32 (0.06, 1.7) | 10.0 (6.7, 41.6) | 47.3 (30.4, 118) | 219 (45, 1186) | 40 (10, 80) |
| Vascular disease | | p=0.155 | p=0.056 | p=0.144 | p=0.2691 | p=0.056 | p=0.156 |
| | 4 | 46.6 (22.6, 70.6) | 1.4 (0.63, 3.3) | 22.6 (10.2, 33.3) | 58.8 (27.6, 94.2) | 790 (386.5, 1801) | 80 (40.5, 360) |
| Asthma | | p=0.382 | p=0.755 | p=0.443 | p=0.106 | p=0.512 | p=0.813 |
| | 22 | 13.1 (5.0, 38.0) | 0.29 (0.06, 0.82) | 4.4 (1.1, 14.2) | 23.7 (6.5, 38.4) | 49.5 (14, 353) | 15 (10, 40) |
| Immunosuppressive | | p=0.376 | p=0.512 | p=0.338 | p=0.835 | p=0.412 | p=0.372 |
| medication | | 8.0 (5.2, 32.0) | 0.12 (0.07, 0.73) | 2.8 (1.2, 11.8) | 37.4 (18.8, 44.4) | 28 (15, 279) | 10 (1, 20) |
| Inhaled steroid | | p=0.112 | **p=0.018** | p=0.101 | p=0.935 | p=0.325 | p=0.050 |
| | 25 | 33.2 (8.0, 43.3) | 0.47 (0.24, 1.03) | 9.2 (2.9, 14.9) | 33.0 (16.1, 45.7) | 203 (34, 310) | 20 (10, 80) |

[A] Values are area under the curve x$10^5$

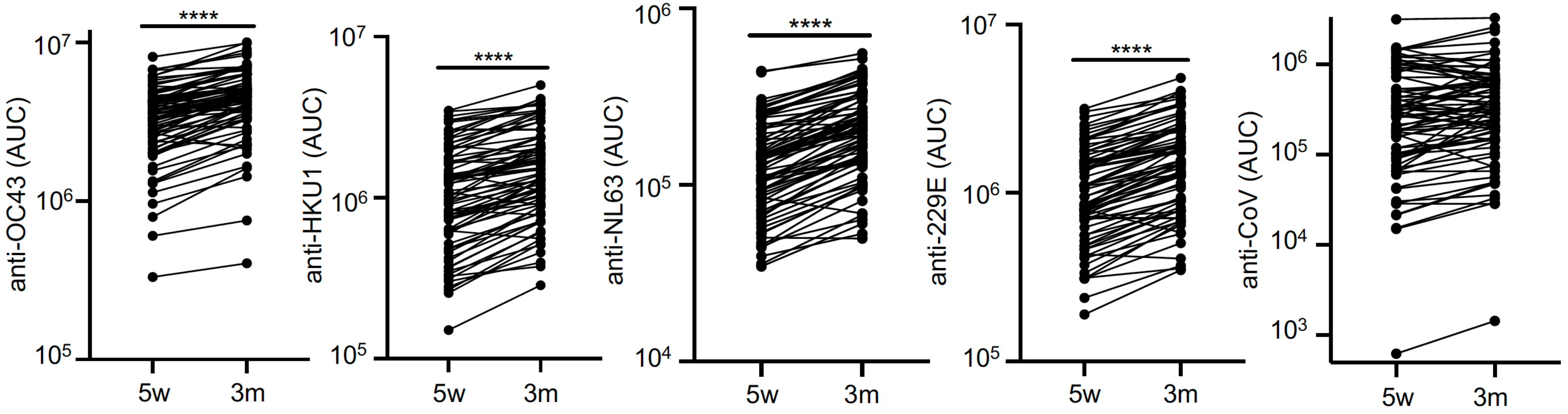


**Supplementary Figure 2. Antibodies to seasonal coronaviruses and SARS-CoV do not decline from five weeks to three months post COVID-19 symptom resolution.** Sera from COVID-19 convalescent subjects five weeks (w) and three months (m) post symptom resolution were evaluated for IgG (reported as AUC) that bound to the spike protein of common cold coronaviruses (CCoV) OC43, HKU1, NL63 and 229E and SARS-CoV by multiplex assay. Lines connect time points for individual subjects (n=79). Paired t tests were performed ****$p<0.0001$.
